# Exploring Microbiota-Associated Metabolites in Twins Discordant for Type 1 Diabetes

**DOI:** 10.1101/2025.02.20.25322611

**Authors:** Elizabeth R. Flammer, Michael W. Christopher, Esabella R. Powers, Hali Broncucia, Andrea K. Steck, Stephen E. Gitelman, Timothy J. Garrett, Heba M. Ismail

## Abstract

**Objective:** Identify microbial and microbiota-associated metabolites in monozygotic (MZ) and dizygotic (DZ) twins discordant for type 1 diabetes (T1D) to gain insight into potential environmental factors that may influence T1D.

**Research Design and Methods:** Serum samples from 39 twins discordant for T1D were analyzed using a semi-targeted metabolomics approach via liquid chromatography-high-resolution tandem mass spectrometry (LC-HRMS/MS). Statistical analyses identified significant metabolites (p < 0.1) within three groups: All twins (combined group), MZ twins, and DZ twins.

**Results:** Thirteen metabolites were identified as significant. 3-indoxyl sulfate and 5-hydroxyindole were significantly reduced in T1D individuals across all groups. Carnitine was reduced, and threonine, muramic acid, and 2-oxobutyric acid were significantly elevated in both All and MZ groups. Allantoin was significantly reduced and 3-methylhistidine was significantly elevated in All and DZ groups.

**Conclusions:** Metabolite dysregulation associated with gut dysbiosis was observed. However, further validation of our findings in a larger cohort is needed.

**Article Highlights:**

- Why did we undertake this study? We believed this cohort of twins discordant for type 1 diabetes (T1D) would allow for control over genetic variability to examine environmental factors.
- What is the specific question(s) we wanted to answer? We aimed to identify differences in microbial and microbiota-associated metabolites in twins discordant for T1D to examine the effect of the gut microbiome on T1D.
- What did we find? Thirteen metabolites were identified as significantly different.
- What are the implications of our findings? Our results show the dysregulation of several microbial metabolites in twin pairs, suggesting that the gut microbiome plays a role in the pathogenesis of T1D.

## MAIN TEXT

Type 1 diabetes (T1D) is an autoimmune disease where T cells attack and destroy insulin-producing beta cells, leading to hyperglycemia and insulin dependency (1). Although T1D is a heritable polygenic disease, most individuals diagnosed with T1D do not have a relative with the disease (2). Monozygotic twins share 100% of their genes, yet the discordant rates of T1D range between 40% and 60%, indicating that environmental factors may contribute (3). Evidence suggests that the gut microbiome may be partially responsible for the immune responses during the progression to T1D (1). The composition of the gut microbiome affects the development and function of the immune system, and their microbial metabolites can mediate immune responses and protective functions (4) by influencing T cell activation and differentiation into helper or regulatory T cells (5). Building on our understanding of the potential influence of gut microbial metabolites on T1D development, we aimed to identify differences in these metabolites in a unique cohort of monozygotic (MZ) and dizygotic (DZ) twins discordant for T1D. This allows for greater control over genetic variability while focusing on microbial factors. Our goal was to identify differences in microbial metabolites across the entire population and within MZ and DZ subsets. Using this cohort, we conducted a semi-targeted metabolomics analysis using liquid chromatography-high-resolution tandem mass spectrometry (LC-HRMS/MS).

### Research Design and Methods

#### Design

Our study consisted of a cohort of 39 twins discordant for T1D: 9 MZ pairs and 10 DZ pairs. The twins are enrolled in the Twin Family Study of Islet Cell Autoimmunity at the Barbara Davis Center for Diabetes. Cotwin subjects were enrolled after the initial proband was diagnosed with T1D and followed for development of islet autoimmunity and/or T1D (6). The serum samples from twin pairs were collected within a three-month window to minimize variation with each pair being discordant for T1D (ie proband with T1D and cotwin without diabetes). The full cohort’s serum collection spanned over the prior eight years, with a focus on local twin populations. Subjects were enrolled with informed consent, and the study was approved by the University of Colorado Institutional Review Board.

#### Analysis

Sample preparation followed a protein precipitation protocol using an extraction solution of 8/1/1 (v/v/v) acetonitrile/methanol/acetone. LC-HRMS/MS data acquisition was performed using a Vanquish LC system coupled to an Orbitrap Exploris 120 (ThermoScientific). Additional information is provided in the Supplemental Materials under ‘Sample Preparation’ and ‘LC-HRMS/MS Instrumentation’.

#### Data Processing

Compound Discoverer (ThermoScientific) was used to perform data processing and provide level 2 metabolite ID’s in positive and negative ionization mode. Full details of the workflow are provided in the supplemental information under ‘Compound Discoverer Parameters’.

#### Statistical Analysis

All multivariate statistical analyses were performed using MetaboAnalyst (7), where data was sum-normalized, log-transformed, and auto-scaled. One participant was identified as an outlier in principal component analysis (PCA) and excluded (Supplementary Fig. 1). Data were not corrected for age, as the PCA plot showed no clustering by age (Supplementary Fig. 1). Because of the rarity of the data set and the small number of discordant twins for diabetes, significance was set at 0.1. Given the limited sample size that reduces the statistical power, a p-value of <0.1 was chosen to help us identify metabolites that could potentially show higher significance in a larger study. Three groups were used during the analysis: ‘All,’ included all monozygotic and dizygotic twin pairs discordant for T1D (n=39). The second group, ‘MZ,’ included only monozygotic twin pairs discordant for T1D (n=19), while the third group, ‘DZ,’ included only dizygotic twin pairs discordant for T1D (n=20). The ‘All’ group aimed to increase statistical power, at the cost of zygosity stratification, and identify metabolites with broader significance, which could then be evaluated within the ‘MZ’ and ‘DZ’ groups in larger future studies. Figures were created in R Studio using R programming language.

### Data and Resource Availability

The data sets generated and analyzed are available from the corresponding author upon reasonable request.

## Results

#Table 1 shows demographic information.

**Table 1.**
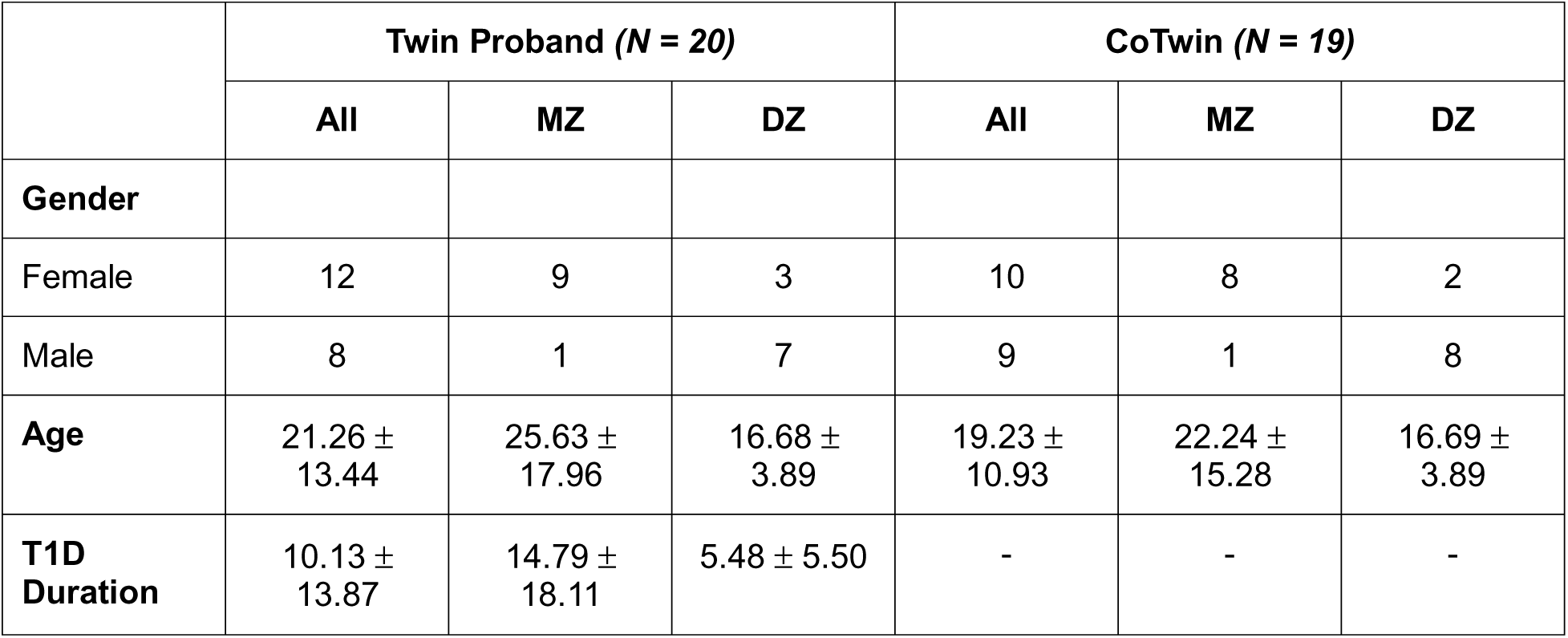
Demographic characteristics of study participants.

Thirteen significant microbial and microbiota-associated metabolites were identified across all three groups. The corresponding p-values, ionization mode (positive (+ve) or negative (−ve)), directional changes of metabolites, and metabolite classification are presented in Table 2.

**Table 2.** Significant metabolites and corresponding p-values associated with sample groupings, reduced or elevated in the T1D cohort. The ionization mode used for detection is denoted by positive (+) or negative (−)

| Metabolite | All | MZ | DZ | Elevation Status (T1D) | Classification |
| --- | --- | --- | --- | --- | --- |
|  | P Value | P Value | P Value |  |  |
| (-) 3-Indoxyl Sulfate | 0.003 | 0.02 | 0.07 | Reduced | Tryptophan Metabolism |
| (+) 5-Hydroxyindole | 0.002 | 0.01 | 0.05 | Reduced | Indole Derivatives |
| (+) Carnitine | 0.01 | 0.02 | - | Reduced | Fatty Acid Oxidation |
| (-) Allantoin | 0.02 | - | 0.04 | Reduced | Purine Metabolism |
| (+) Allantoin | 0.02 | - | 0.04 | Reduced | Purine Metabolism |
| (+) Dihydrothymine | 0.08 | - | - | Reduced | Pyrimidine Metabolism |
| (+) Uric Acid | 0.07 | - | - | Reduced | Purine Metabolism |
| (+) Betaine | - | 0.06 | - | Reduced | Methyl Donor |
| (+) 2-Hexenoylcarnitine | - | - | 0.08 | Reduced | Fatty Acid Oxidation |
| (+) Threonine | 0.04 | 0.06 | - | Elevated | Amino Acid Metabolism |
| (+) Muramic Acid | 0.03 | 0.08 | - | Elevated | Bacterial Metabolism |
| (-) 2-Oxobutyric Acid | 0.04 | 0.07 | - | Elevated | Amino Acid Metabolism |
| (+) 3-Methylhistidine | 0.05 | - | 0.02 | Elevated | Histidine Metabolism |
| (+) Hippuric Acid | 0.07 | - | - | Elevated | Bacterial Metabolism |

Within the All group in −ve mode, 3-indoxyl sulfate and allantoin (P <0.1, and P <0.05, respectively) were reduced in T1D twins, while 2-oxobutyric acid (P < 0.1) was elevated. In +ve mode, 5-hydroxyindole, carnitine, allantoin, dihydrothymine, uric acid, and 2-hexonylcarnitine (P <0.05, P <0.05, P <0.05, P <0.1, P <0.1, and P <0.1, respectively) were reduced, whereas threonine, muramic acid, 3-methylhistidine and hippuric acid (P <0.1, P <0.1, P <0.05, and P <0.1, respectively) showed elevated levels in individuals with T1D.

Among these, 3-indoxyl sulfate and 5-hydroxyindole were significant across all groups (Fig. 1).

**Figure 1:**
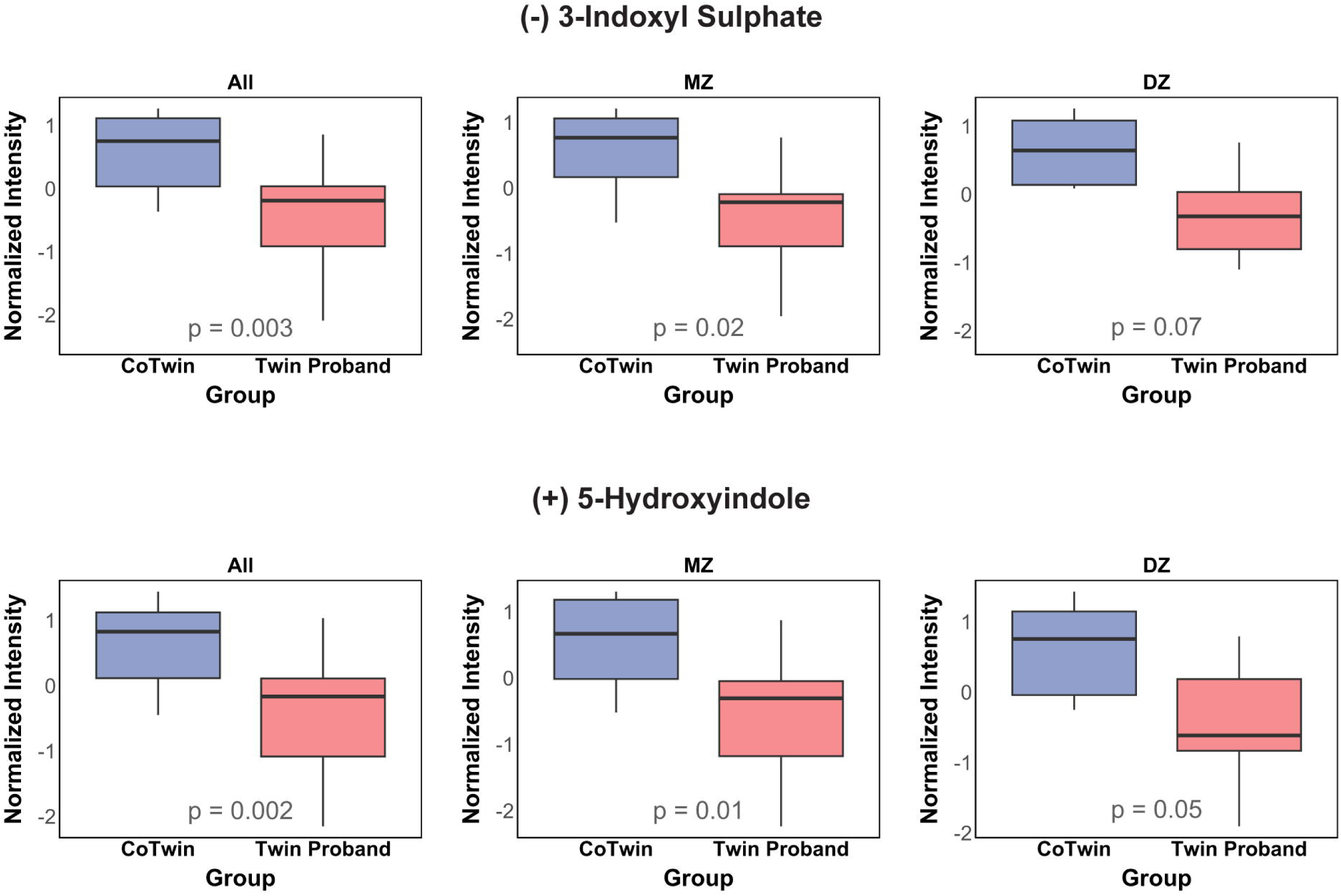
Boxplots depicting differences in 3-indoxyl sulfate and 5-hydroxyindole across all three groups.

In both the All and MZ groups, carnitine, threonine, muramic acid, and 2-oxobutyric acid were found to be significant. Allantoin and 3-methylhistidine were significant in both All and DZ groups. Within the MZ group, betaine was significant, while dihydrothymine, hippuric acid, and uric acid were only significant in the All group. Finally, 2-hexenoylcarnitine was significant in the DZ group (Fig. 2).

**Figure 2:**
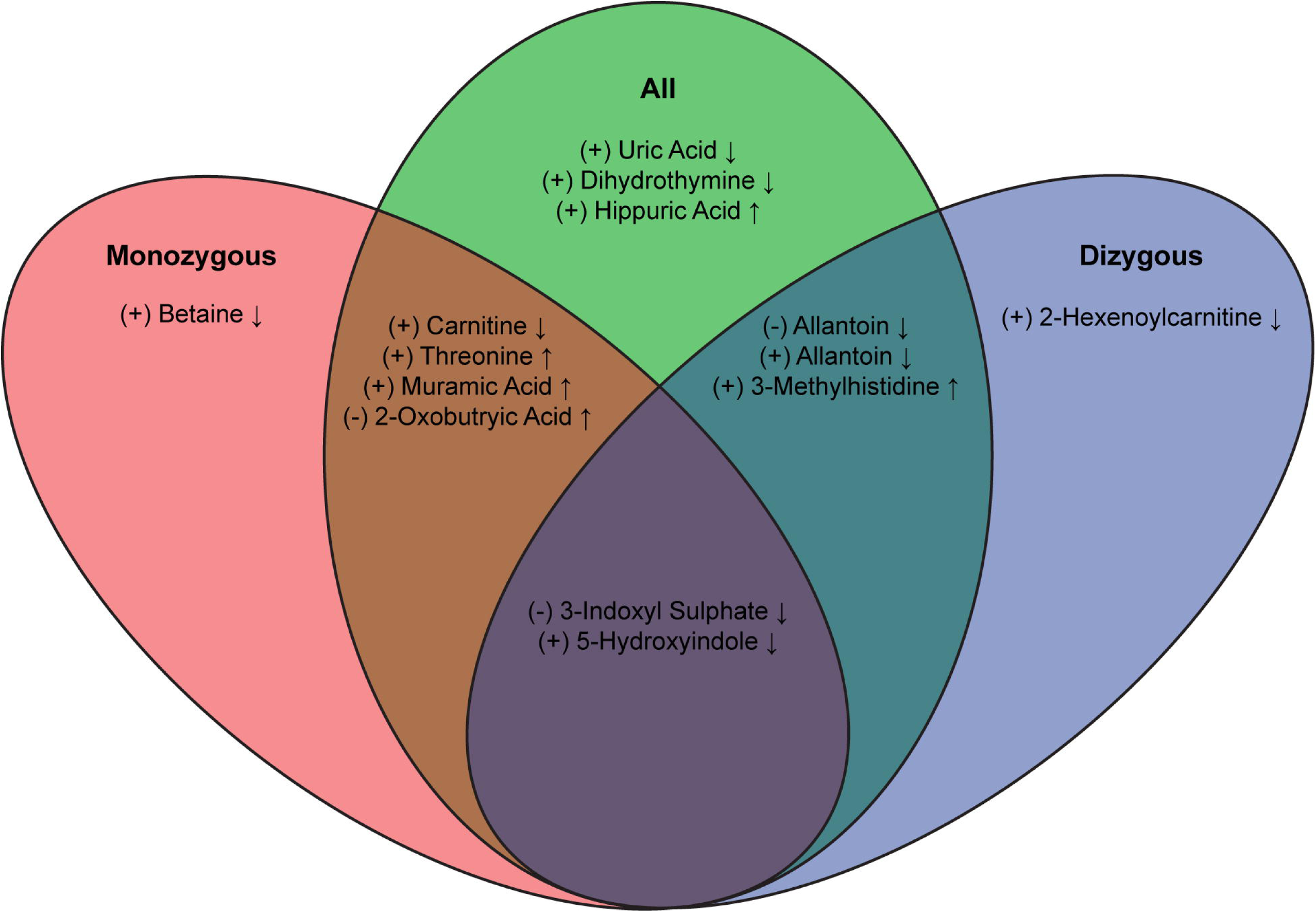
Venn diagram illustrating the overlap of significant metabolites across all three groups, depicting shared and group-specific significant metabolites.

## Conclusions

Using this unique sample population, we aimed to examine the possible effects of the gut microbiome on disease pathogenesis through the identification of statistically significant microbiota-associated metabolites. This study identified thirteen significant metabolites across all three groups. Several of these metabolites, including 3-indoxyl sulfate, 5-hydroxyindole, muramic acid, and hippuric acid, have previously been associated with gut dysbiosis (8,9,10).

Among identified metabolites, reduced levels of 3-indoxyl sulfate and 5-hydroxyindole, byproducts of tryptophan metabolism produced by gut bacteria, in individuals with T1D demonstrate a link between microbiome disruption and T1D. Reduced levels of 3-indoxyl sulfate and 5-hydroxyindole have been associated with gut dysbiosis and increased inflammation (8,11). Muramic acid, a component of peptidoglycan, could indicate increased microbial exposure when found at elevated levels (9). Interestingly, hippuric acid was elevated despite previous literature correlating increased levels with increased gut microbiome diversity (10).

Dysregulations in amino acids and their derivatives were also observed. Amino acids can be produced by both microbial and host sources since both require them for normal function, and changes in amino acids affect the gut microbiome. Carnitine, a quaternary ammonium compound, although not synthesized by bacteria, is utilized by gut microbes. Both carnitine and acylcarnitine are involved in the endogenous metabolism of short-chain fatty acids and contribute to maintaining a healthy gut microbiota (12). In this study, reduced levels of carnitine and 2-hexenoylcarnitine, a specific acylcarnitine, were observed. These reductions have been previously reported in T1D (13) and may contribute to dysregulation of short-chain fatty acids and the gut microbiome (12). Similarly, gut microbiota can contribute to the production of betaine, which can positively influence gut microbiome composition. Reduced betaine levels have been previously associated with decreased insulin sensitivity in patients with type 2 diabetes (14). Additionally, 3-methylhistidine is a posttranslational modification that can be orchestrated by microbes. These modifications can contribute to the production of autoantigens that stimulate T cells, leading to autoimmunity (15). Elevated levels of threonine and 2-oxobutryic acid, its degradation product, have been associated with T1D and a lack of endogenous insulin (16,17).

Several metabolites, including dihydrothymine, uric acid, and allantoin, were found to be reduced in this study. The exact mechanisms responsible for these changes are not fully understood. In our study, we found reduced levels of uric acid which is normally associated with improved gut health and lowered inflammation (18). Uric acid is converted to allantoin, and the reduced levels of allantoin observed are likely a consequence of decreased uric acid levels (18).

A limitation of our study is the small sample population, which may affect the generalizability of our findings. Expanding the cohort in future studies will be essential for enhancing the robustness of our analysis. Notably, twins account for approximately 3% of the US population and instances where one twin is diagnosed with T1D while the other remains unaffected are uncommon. This necessitated a small-scale initial study. Additionally, some of the changes could be related to glycemic changes in those probands with T1D. However, measures of glycemia or HbA1c were unavailable to us.

Strengths include the analysis of a unique cohort of local monozygotic and dizygotic twins followed longitudinally at the Barbara Davis Center. This reduces environmental differences that may affect metabolite variations unrelated to T1D status. Moreover, LC-HRMS/MS is a powerful tool that allows for the detection of subtle differences in metabolite profiles, crucial for studying twin pairs and diseases such as T1D.

In conclusion, our results show the dysregulation of microbiota-associated metabolites, as observed in twins discordant for T1D, suggesting that the gut microbiota play a role in T1D pathogenesis. Further research is necessary to determine the specific roles of these metabolites, regulatory mechanisms, and potential contributions to T1D. Moreover, additional studies are needed to understand why certain metabolites are dysregulated in specific zygotic groups. These findings could ultimately result in a novel means of identifying individuals at risk for T1D.

## Data Availability

All data produced in the present study are available upon reasonable request to the authors.

## ACKNOWLEDGMENTS

**Funding and Assistance.** This study was supported by the Nation Institute of Diabetes and Digestive and Kidney Diseases of the Nation Institutes of Health (DK-129799) **Duality of Interest.** No potential conflicts of interest relevant to this article were reported.

## Author Contributions

E.R.F. performed sample analysis, data analysis and interpretation, and wrote the manuscript. M.W.C. and E.R.P. aided in data acquisition and interpretation and edited the manuscript. H.B. and A.K.S. provided samples, reviewed findings, and edited the manuscript. S.E.G. reviewed findings and edited the manuscript. H.M.I. and T.J.G. provided oversight of the research, reviewed all data and findings, and edited the manuscript. T.J.G. is the guarantor of this work and, as such, had full access to all the data in the study and takes responsibility for the integrity of the data and the accuracy of the data analysis.

## Prior Presentation

Parts of this study were presented as a poster at the 72nd Annual American Society for Mass Spectrometry Conference, June 2024, Anaheim, CA.

---

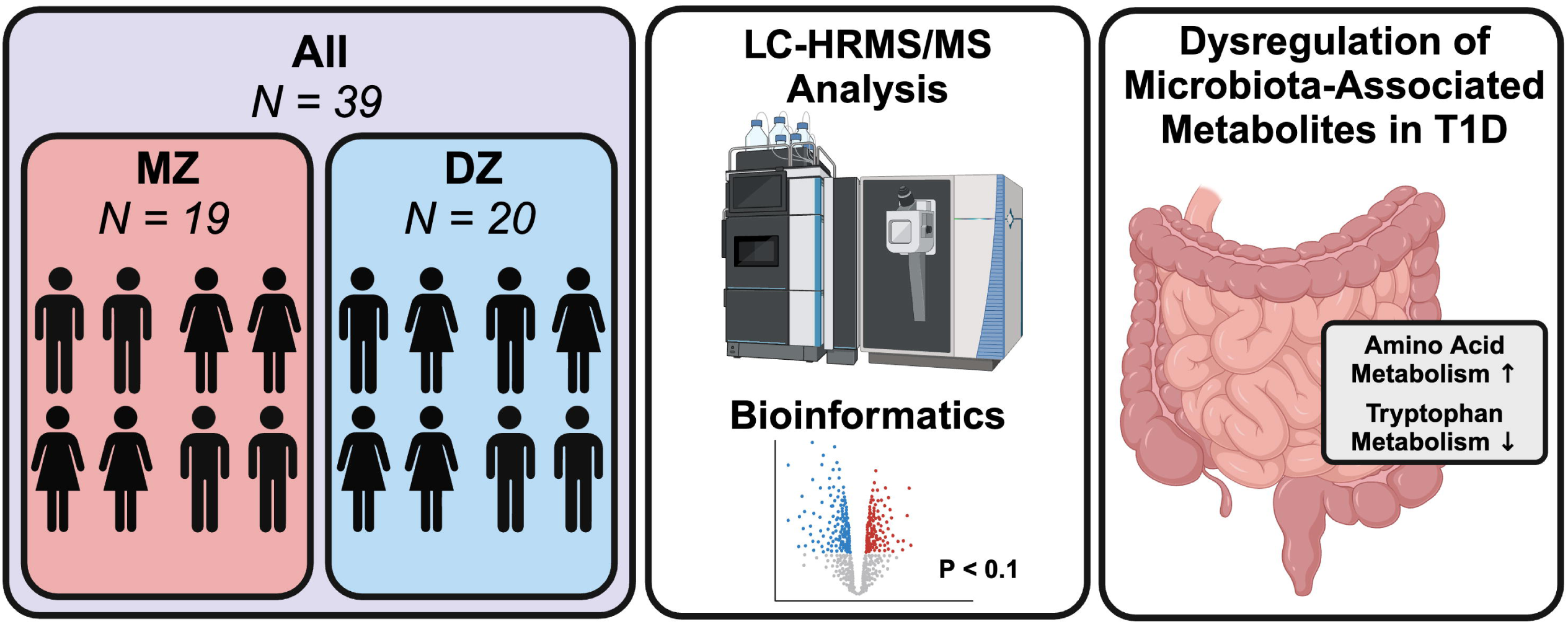

## Notes

### Competing Interest Statement

The authors have declared no competing interest.

### Author Declarations

The study was approved by the University of Colorado Institutional Review Board.

